# Addressing a complicated problem: can COVID-19 asymptomatic cases be detected – and epidemics stopped− when testing is limited and the location of such cases unknown?

**DOI:** 10.1101/2020.11.10.20223495

**Authors:** Ariel L. Rivas, Almira L. Hoogesteijn, James B. Hittner, Folorunso O. Fasina, Marc H.V. van Regenmortel

## Abstract

Can the COVID-19 pandemic be stopped when the principal disseminators −asymptomatic cases− are not easily observable? This question was addressed exploring the cumulative epidemiologic data reported by 51 countries, up to October 2, 2020. In particular, the validity of test positivity and its inverse (the ratio of tests performed per case detected) to indicate whether asymptomatic cases are being detected and isolated –even when only a minor percentage of the population is tested− was evaluated. By linking test positivity data to the number of COVID-19 related deaths reported per million inhabitants, the research question was answered: countries that expressed a high percentage of test positivity (>5%) reported, on average, 15 times more deaths than countries that exhibited <1% test positivity. It is suggested that such a large difference in outcomes is due to the exponential growth that epidemics may experience when silent (asymptomatic) cases are not detected and, consequently, the disease disseminates. Because temporal and geo-referenced data on test positivity may facilitate cost-effective, site-specific testing policies, it is postulated that the risk of uncontrolled epidemics may be ameliorated when test positivity is investigated.

## Introduction

The prompt identification and isolation of COVID-19 asymptomatic cases is a critical problem that requires an immediate solution. Analyzing data from dozens of countries^**1**^ and adopting an interdisciplinary approach^**2**^, here we explore whether asymptomatic cases can be detected rapidly and cost-effectively, even when testing is limited, and link the findings to economic data recently released.

The potential exponential growth of epidemics is a composite problem that includes four elements. The first one is the fact that asymptomatic cases are the principal spreaders of this pandemic. Because they do not reveal symptoms, these patients do not seek medical assistance and can disseminate the virus silently.^**3**^ To detect asymptomatic cases, the theoretical solution is to test all citizens.

Unfortunately, there is a second problem: no country can currently test, daily, 100% of its population. While remarkable progress towards universal and simultaneous testing has recently been achieved, usually, less than 4/1000 people are tested/day.^**4, 5**^

These two problems converge, creating a third problem (a vicious cycle): how to identify the optimal test that determines disease prevalence (which, in epidemics, may rapidly change) given the fact that disease prevalence influences test sensitivity and specificity and a test with high sensitivity and specificity is needed to determine disease prevalence.^**6**^ The circular nature of this problem is aggravated by COVID 19-related limited testing, which prevents the estimation of disease prevalence. Consequently, no specific test is investigated here. Instead, a previous (and more urgent) question is addressed: what testing metric is more adequate to estimate disease prevalence? This question matters because, when disease prevalence is shown to decrease over time, it would indicate that asymptomatic cases are being detected.

In addition, economics –the fourth dimension of the composite problem− should also be considered. Economic aspects involve, at least, two dimensions (i) cost-effective approaches that can be rapidly implemented even when testing is very limited, and (ii) connections between public health and the economy at large.

These four challenges were simultaneously investigated by assessing biological-geographical-temporal-economic relationships described by the Test Positivity percentage (TP %) and its inverse (the Tests/Case [T/C] ratio). The TP % is the proportion of tested individuals that yield virus-positive results (number of cases / number of tests x 100). The T/C is the ratio between the number of tests conducted per each case detected. Given that asymptomatic cases, when undetected, spread the virus, larger numbers of infections and deaths are expected (disease prevalence will increase) when testing shows high TP percentages or low T/C ratios. Accordingly, here it is explored whether the problems associated with asymptomatic case-driven epidemic growth, as well as the interdependence between disease prevalence and test evaluation, and the limited resources (including limited testing) frequently reported in epidemic control campaigns can be circumvented by measuring metrics that, indirectly, estimate disease prevalence and its main drivers –asymptomatic cases.

## Materials and methods

COVID-19 census data available in the public domain (Supplementary material - epidemiologic and economic data on COVID-19 reported up to October 2, 2020) were descriptively analyzed using a commercial package (Minitab 19, State College, PA, Minitab Inc,). Specifically, the measure of central tendencies were obtained through the population medians using the non-parametric statistical procedure, the Mann-Whitney test. They included (i) cumulative epidemiologic data from 51 countries (*Worldometer*, https://www.worldometers.info/coronavirus/ ; and *Ourworldindata*, https://ourworldindata.org/coronavirus-testing#how-many-tests-are-performed-each-day) and (ii) economic data from 28 countries reported by *Ourworldindata*, which does not provide updated (2020) GDP data for all countries (https://ourworldindata.org/covid-health-economy, accessed October 2, 2020). Table 1 (Supporting materials) reports the data analyzed.

This selected dataset met several criteria: (i) to include countries reporting their first COVID-19 cases at or before April 15, 2020, so outcomes would reflect epidemic processes five or more months long; (ii) to demonstrate geo-demographical diversity (small and large areas, small and large populations, coastal and non-coastal countries); and (iii) to possess socio-economic diversity (countries with various levels of socio-economic development). The purpose of the inclusion criteria was not to pursue representativity but plausibility so that, if similar outcomes were observed in countries that differed geo-demographically and/or economically, findings would not be attributed to such conditions.

In addition, a small geo-referenced and temporal dataset was examined, which included the county-level test positivity data reported by the Puerto Rico Public Health Trust on Sept 18 and 21, 2020 (https://experience.arcgis.com/experience/fe19b3ab2a9c4fcd952c27686fe136c4). Municipality-level demographic and surface data were extracted from the US Census Bureau and Puerto Rico Administrative Division (https://www.census.gov/data/tables/time-series/demo/popest/2010s-total-puerto-rico-municipios.html and https://www.citypopulation.de/en/puertorico/admin/, accessed Nov 8, 2020)

## Results

When the test positivity percentage and the ratio between tests performed per case detected were explored with all the (cumulative) data generated by 51 countries up to Oct 2, 2020, an orthogonal distribution was observed (Fig. **1A**). Demonstrating its validity to identify asymptomatic cases, the TP % showed a positive and statistically significant correlation with deaths/million inhabitants (deaths/mi, Fig. **1B**). Because the T/C ratio displayed an L-shaped distribution, the T/C ratio was a better discriminator than the TP%: without cutoffs, the T/C ratio differentiated two groups of countries (A and B), which displayed non-overlapping values of deaths/mi (a third group of countries [C] displayed values that partially overlapped with the remaining groups [Fig. **1C**]). Countries clustered into the B group exhibited a median number of deaths/mi 15 times higher than A countries (*p*<0.01, Mann-Whitney test, Fig. **1D**). Therefore, the T/C ratio also demonstrated its validity. When the Gross Domestic Product (GDP) change reported in 28 countries in the second quarter of 2020 was considered^**1**^, a statistically significant correlation was found in relation to both the TP % and the T/C ratio (Figs. **1 E, F**).

**Fig. 1.**
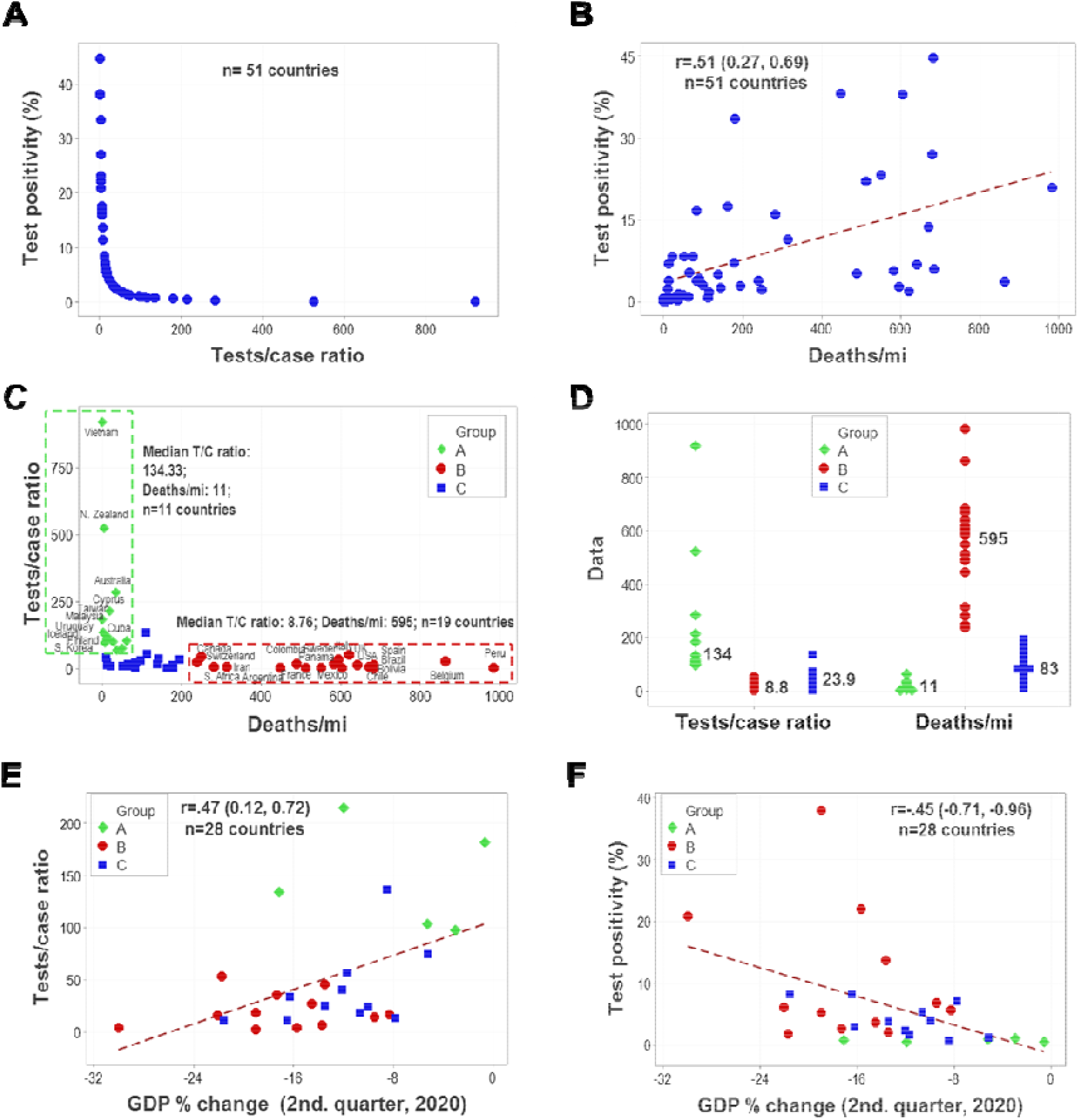
Detecting the principal disseminators of the pandemic, even with limited testing. The percentage of test positivity (TP %) and its inverse (the tests/case ratio) were investigated in 51 countries with all the cumulative data reported up to October 2, 2020. When both metrics were simultaneously assessed, an orthogonal data distribution was observed (**A**). The percentage of test positivity was positively and significantly correlated with deaths/million inhabitants (r=.51, *p*< 0.001, **B**). Such a finding suggests that, when the TP percentage is high, symptomatic cases are predominantly identified −those usually tested when they seek medical assistance, because they feel ill. When asymptomatic cases are missed (when TP % is high or the T/C ratio is low), the epidemic disseminates and may grow exponentially. Such a hypothesis was supported when the T/C ratio was explored together with the number of deaths reported per million inhabitants (deaths/mi): the T/C ratio was orthogonal with respect to deaths/mi –a pattern that differentiated three groups of countries, of which two showed non-overlapping data distributions (**C**). Supporting the hypothesis that high T/C values are associated with low numbers of deaths/mi (because asymptomatic cases are detected and isolated, which prevents the exponential growth of epidemics), countries clustered in the A group reported, on average (median), 134 tests per each case detected as well as 11 deaths/mi. In contrast, B countries (which had performed 8.8 tests per case detected) reported a median of 595 deaths/mi (*p*<0.01, Mann-Whitney test, **D**). The TP % and the T/C ratio also demonstrated significant correlations with the economy. While only 28 of the 51 original countries were investigated (no updated data for the second quarter of 2020 was available for the remaining countries), the change in the Gross Domestic Product was significantly correlated with the TP % and the T/C ratio (**E, F**). The countries clustered in these groups are: (group A): Australia, Cuba, Cyprus, Finland, Iceland, Malaysia, New Zealand, S. Korea, Taiwan, Uruguay, and Vietnam; (group B): Argentina, Belgium, Bolivia, Brazil, Canada, Colombia, Chile, France, Iran, Italy, Mexico, Panama, Peru, S. Africa, Spain, Sweden, Switzerland, UK, and USA; and (group C): Belarus, C. Rica, Czechia, Denmark, Germany, Greece, Hungary, India, Israel, Japan, Kenya, Kyrgyzstan, Libya, Norway, Philippines, Portugal, Russia, Saudi Arabia, Slovakia, Tunisia, and Turkey.

Because these metrics can be analyzed with geo-referenced data, cost-benefit oriented decisions can rapidly be planned and deployed (Figs. **2 A, B**). Vice versa, lack of prompt, geo-epidemiologically specific interventions can rapidly induce epidemic growth, which (as illustrated in one geo-temporal specific example) may cover ∼ 7 times more territory and put under risk ∼ 50% more people within 3 days even when the level of testing remained unchanged (Figs. **2 A, B**).

Additional analyses differentiated between cutoff-based, prediction-oriented and pattern recognition-based, real-time, interdisciplinary analysis. Exploring the policy announced by WHO – which assumed that countries reporting less than 5% TP could regard the epidemic to be under control^**7**^−, 51 countries were divided into three groups according to their TP values (<1%, 1−5%, or >5% TP, Fig. **3A**). Findings revealed a substantial number of deaths even below the 5% cutoff mentioned by WHO (Figs. **3B-D**). The hypothesis that a cutoff could predict the imminent end of an epidemic was further negated when the number of tests performed per million inhabitants (tests/mi) was considered (Fig. **3E**). No statistically significant correlation was found between the number of tests performed and COVID-19 related deaths (Fig. **3F**).

**Fig. 2.**
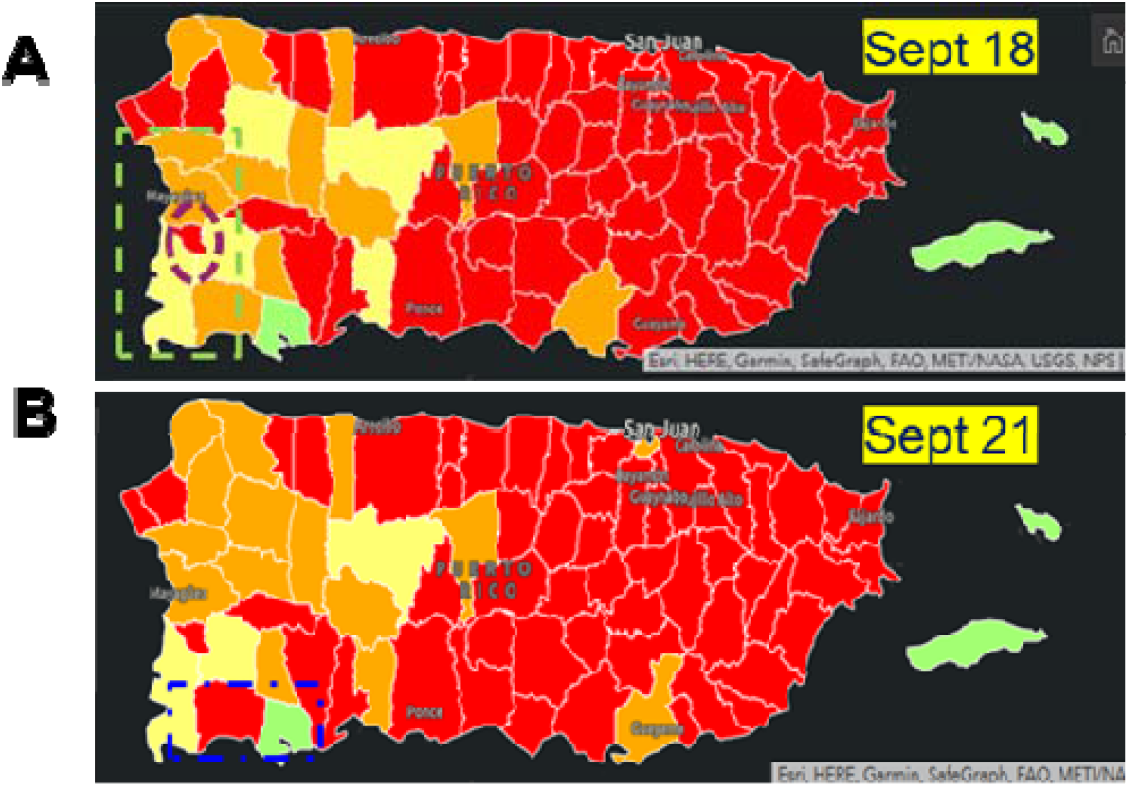
Geo-temporal, cost-benefit, real-time analysis and intervention. The daily test positivity (TP) percentages of all Puerto Rican municipalities is shown as reported by the Puerto Rico Public Health Trust on Sept 18 and 21, 2020 (https://experience.arcgis.com/experience/fe19b3ab2a9c4fcd952c27686fe136c4). Colors denote TP percentage: < 3% (green), 3.00-5.99 (yellow), 6-9.99 (orange), or >10% (red). The Hormigueros municipality (red area within the purple oval, **A**) occupies 29 sq km, which represents 3.39% of the area (853.49 sq km) and hosts 17,250 inhabitants (or 6.48%) of the population (266,265 inhabitants) depicted in the green rectangle (**A**). If all the resources invested in regional testing were equal to 100 units (some measure that would include diagnostic kits, human and other resources) and they were proportionally distributed across municipalities, then the Hormigueros municipality would receive 6.48 units. If –considering its high TP %− testing was increased 500% in Hormigueros (receiving 32.4 of the 100 units of resources assigned to the region), such a policy would reduce the testing corresponding to the remaining municipalities in, approximately, 27 % (from the original 93.52 units [100-6.48], to 67.6 [100-32.4], or 67.6/93.52= 72.28 % of the original resource units). This means that, without changing the overall budget or resources assigned to the region, a differential testing policy that increased five times the intervention of a small area displaying high TP could be implemented with a marginal reduction (27%) in the remaining municipalities of the region. If such a policy resulted in a reduction of the TP corresponding to the Hormigueros municipality, the whole region would benefit without increasing any cost. Later data may be considered to estimate the impact of an intervention not implemented. The Lajas municipality (199 sq km, 25,753 inhabitants) displayed, 3 days later, a higher (>10 %) TP (the red area shown within the blue rectangle, **B**). While other explanations cannot be ruled out, one possible cause of the higher TP observed in Lajas is the absence of an intervention implemented in the Hormigueros municipality, which facilitated an epidemic spread that increased 6.86 times in area (from 29 to 199 sq km) and put almost 50% (25,753/17,250) more people at risk even though the testing effort remained unchanged (**B**).

**Fig. 3.**
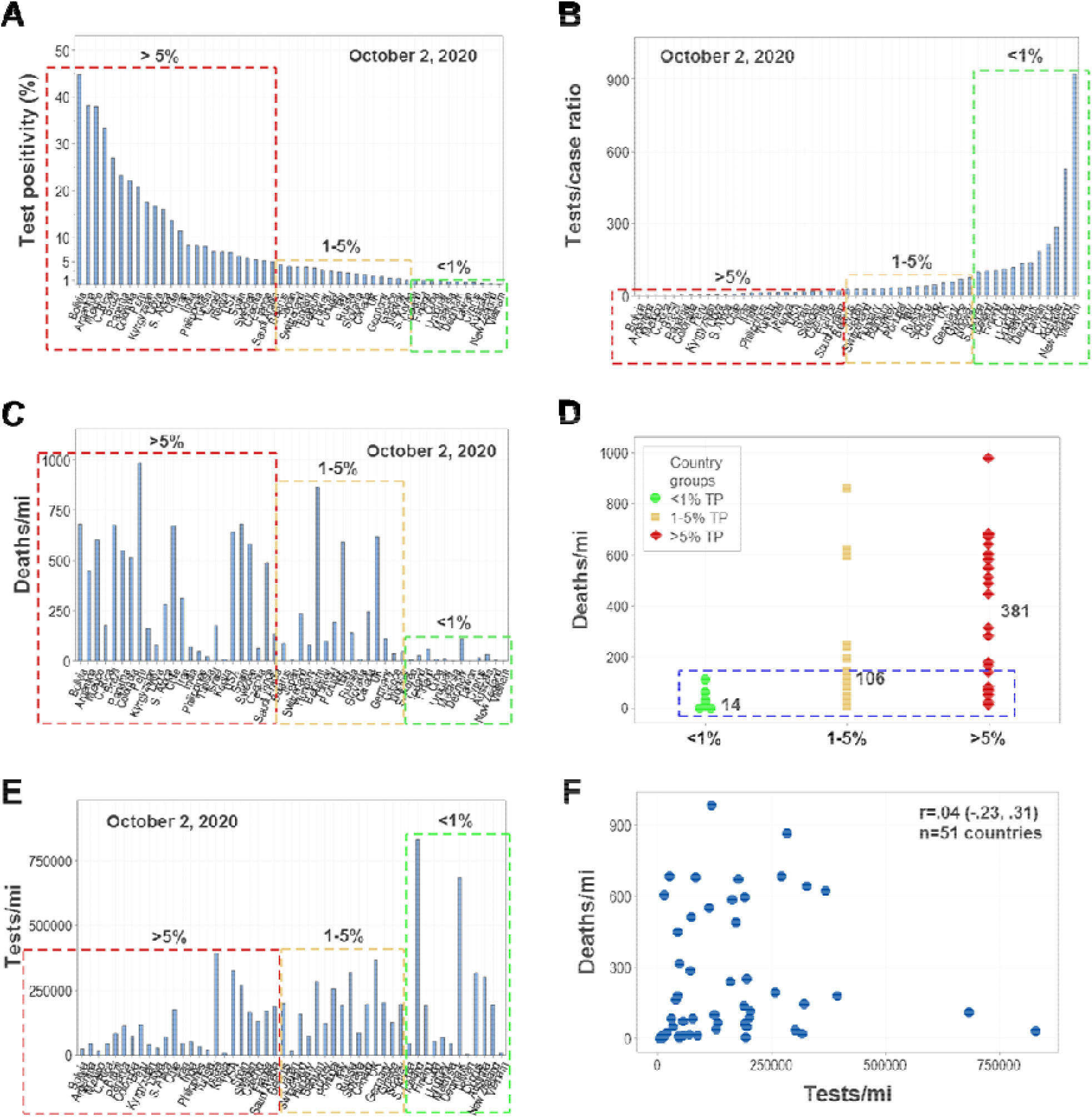
Pattern recognition- vs. cutoff-based discrimination. To elucidate whether a cutoff (TP <5 %) identifies epidemics under control, several analyses based on numerical thresholds were conducted. First, 51 countries were divided into three groups, according to their TP % (<1%, 1−5%, or >5% TP, **A**). Using the same classification, the T/C ratio and the number of deaths reported per million inhabitants (deaths/mi) were assessed (**B, C**). Findings showed that in both the 1−5% and the >5% TP groups, high numbers of deaths/mi were reported in numerous countries (**C**). Across cutoff-based groups, a substantial number of countries displayed overlapping numbers of deaths/mi (blue rectangle, **D**). Findings did not depend on the metric used: both percentages (such as the TP), ratios (e.g., the T/C ratio) and counts (number of tests performed per million inhabitants) did not show patterns when cutoffs were considered (**A-E**). Furthermore, count data were not informative: the correlation between the number of tests performed and the number of COVID-19 related deaths (adjusted per million inhabitants) was not statistically significant (**F**).

## Discussion

Findings indicated that, alone, no cutoff-based assessment −whether related to a percentage, as in the TP %; a ratio (the T/C index); or a count, as in the number of tests performed/mi− could accurately and consistently inform on epidemic progression. In addition, information on geo-epidemiological relationships was also needed.

Because they reveal data overlapping and lose information, cutoff-based analyses are errorprone.^**8**^ Here the use of cutoff-based assessments was rejected three times. In contrast, cutoff-free metrics (the TP % and the T/C ratio) fostered cost-benefit oriented decisions, rapidly, at specific geographical sites, using numbers but not depending on numerical thresholds. Unlike studies that claim to predict the future −by definition, untestable assumptions or theories−, an approach that measured actual (not assumed) patterns, recognized informative patterns, in real-time. Because the TP % and T/C ratio identified asymptomatic cases, they could be used in policies that pursue epidemic control.

Together, findings highlight the relevance of testing that captures asymptomatic patients. Testing efforts limited to symptomatic cases (those likely to seek medical assistance because they feel ill) are likely to fail because, when an epidemic is under an exponential growth process, it will overrun any hospital-centered contingency plan. When COVID-19 cases grow 60,000 times in just 60 days –as observed in some countries, in 2020−, no hospital can increase its number of beds, ventilators, and personnel in similar magnitudes. In contrast, testing aimed at detecting and isolating asymptomatic cases, when grounded on geo-referenced data of high resolution, may succeed.

These metrics also demonstrated connections between public health and the economy. The TP percentages and T/C ratios indicated that countries where asymptomatic cases were not explicitly investigated also experienced uncontrolled epidemic spread, the largest numbers of deaths/mi, and the largest economic declines. Vice versa, countries where the T/C ratio was higher or the TP% was lower –those that emphasized the detection of asymptomatic cases− reported much fewer deaths and the smallest economic declines.

Because epidemics, if not rapidly eradicated, can grow exponentially (generating colossal losses in lives and livelihoods), findings corroborated −now on a worldwide scale− that expenditures in public health are not costs but investments that yield high social returns on the investment (ROI). While previous studies have estimated the ROI of public health at 88 units of benefit per each cost unit, the fact that up to one fifth of the national GDP can be lost within a few weeks when asymptomatic cases are not detected –as shown in figures **1 E** and **F**− suggest that the benefits induced by public health may be higher than previously estimated.^**9-11**^ It is postulated that, to materialize and expand a positive relationship between public health and the economy, current paradigms may need to be re-assessed. While reductionist approaches may be invalid (such as the assumption that continuous data can be dichotomized without inducing errors^**12**^), interdisciplinary approaches may support local and global efforts meant to rapidly and effectively identify and isolate asymptomatic cases and, consequently, prevent the dissemination of COVID-19.

## Supporting information

Epidemiologic and economic data on COVID-19

## Data Availability

All data are publically available.

https://ourworldindata.org/coronavirus-testing#how-many-tests-are-performed-each-day

