## Supplementary material for "Addressing a complicated problem: can COVID-19 asymptomatic cases be detected – and epidemics stopped− when testing is limited and the location of such cases unknown?": Epidemiologic and economic data on COVID-19

**Supplementary materials**

**Table 1. Epidemiologic and economic data on COVID-19 reported up to October 2, 2020**

| **Country** | **Cases^a^** | **Tests^a^** | **TP^b^** | **T/C^c^** | **Deaths^d^** | **Tests^d^** | **T/C-D^e^** | **TP %^f^** | **GDP %^g^** |
| --- | --- | --- | --- | --- | --- | --- | --- | --- | --- |
| Argentina | 765002 | 2002975 | 38.19 | 2.61 | 448.0 | 44216 | B | >5% | -19.0 |
| Australia | 27109 | 7696270 | 0.35 | 283.90 | 35.0 | 300940 | A | <1.0% | -13.5 |
| Belarus | 79421 | 1897263 | 4.18 | 23.88 | 89.0 | 200800 | C | 1-4.99% | * |
| Belgium | 121059 | 3284217 | 3.68 | 27.12 | 864.0 | 283063 | B | 1-4.99% | * |
| Bolivia | 135716 | 303107 | 44.77 | 2.23 | 683.0 | 25878 | B | >5% | * |
| Brazil | 4849229 | 17900000 | 27.09 | 3.69 | 680.0 | 84061 | B | >5% | -8.3 |
| C. Rica | 76828 | 229178 | 33.52 | 2.98 | 180.0 | 44885 | C | >5% | * |
| Canada | 162320 | 7423052 | 2.18 | 45.73 | 249.0 | 196244 | B | 1-4.99% | -8.5 |
| Chile | 466590 | 3394522 | 13.74 | 7.27 | 672.0 | 177189 | B | >5% | -17.3 |
| Colombia | 835339 | 3776280 | 22.12 | 4.52 | 513.0 | 74017 | B | >5% | * |
| Cuba | 5718 | 628725 | 0.90 | 109.95 | 11.0 | 55517 | A | <1.0% | * |
| Cyprus | 1789 | 384821 | 0.46 | 215.10 | 18.0 | 318144 | A | <1.0% | -17.1 |
| Czechia | 76017 | 1410357 | 5.38 | 18.55 | 65.0 | 131636 | C | >5% | -12.1 |
| Denmark | 28932 | 3965265 | 0.72 | 137.05 | 112.0 | 683974 | C | <1.0% | -5.2 |
| Finland | 10244 | 1062584 | 0.96 | 103.72 | 62.0 | 191701 | A | <1.0% | -10.7 |
| France | 577505 | 11020523 | 5.24 | 19.08 | 490.0 | 171097 | B | >5% | -11.9 |
| Germany | 297429 | 16999253 | 1.74 | 57.15 | 114.0 | 202729 | C | 1-4.99% | * |
| Greece | 19346 | 1328042 | 1.45 | 68.64 | 38.0 | 127575 | C | 1-4.99% | * |
| Hungary | 28631 | 740043 | 3.86 | 25.84 | 83.0 | 76656 | C | 1-4.99% | * |
| Iceland | 2809 | 283504 | 0.99 | 100.92 | 29.0 | 829434 | A | <1.0% | * |
| India | 6438968 | 76717728 | 8.39 | 11.91 | 73.0 | 55455 | C | >5% | -11.7 |
| Iran | 464596 | 4067861 | 11.42 | 8.75 | 315.0 | 48276 | B | >5% | -13.5 |
| Israel | 258920 | 3626704 | 7.13 | 14.00 | 178.0 | 394310 | C | >5% | -5.2 |
| Italy | 319908 | 11572459 | 2.76 | 36.17 | 595.0 | 191473 | B | 1-4.99% | * |
| Japan | 84215 | 2125762 | 3.96 | 25.24 | 12.0 | 16821 | C | 1-4.99% | -15.7 |
| Kenya | 38923 | 555711 | 7.00 | 14.27 | 13.0 | 10279 | C | >5% | -7.8 |
| Kyrgyzstan | 47056 | 267718 | 17.57 | 5.68 | 163.0 | 40867 | C | >5% | * |
| Libya | 35717 | 212980 | 16.77 | 5.96 | 83.0 | 30891 | C | >5% | * |
| Malaysia | 11771 | 1581208 | 0.74 | 134.33 | 4.0 | 48698 | A | <1.0% | * |
| Mexico | 748315 | 1968556 | 38.01 | 2.63 | 604.0 | 15228 | B | >5% | -13.7 |
| N. Zealand | 1848 | 970641 | 0.19 | 525.23 | 5.0 | 194047 | A | <1.0% | -21.6 |
| Norway | 14149 | 1065340 | 1.32 | 75.29 | 50.0 | 196125 | C | 1-4.99% | * |
| Panama | 113342 | 488048 | 23.22 | 4.30 | 551.0 | 112669 | B | >5% | -9.5 |
| Peru | 818297 | 3908125 | 20.93 | 4.77 | 983.0 | 118118 | B | >5% | * |
| Philippines | 316678 | 3800150 | 8.33 | 12.00 | 51.0 | 34564 | C | >5% | -21.7 |
| Portugal | 77284 | 2625120 | 2.94 | 33.96 | 195.0 | 257641 | C | 1-4.99% | * |
| Russia | 1194643 | 46823879 | 2.55 | 39.19 | 144.0 | 320820 | C | 1-4.99% | * |
| S. Africa | 676084 | 4209049 | 16.06 | 6.22 | 283.0 | 70745 | B | >5% | -22.1 |
| S. Korea | 23952 | 2333777 | 1.02 | 97.43 | 8.0 | 45510 | A | 1-4.99% | * |
| S. Arabia | 335578 | 6592660 | 5.09 | 19.64 | 138.0 | 188637 | C | >5% | * |
| Slovakia | 11617 | 474845 | 2.44 | 40.87 | 10.0 | 86963 | C | 1-4.99% | -19.0 |
| Spain | 778607 | 12723989 | 6.11 | 16.34 | 684.0 | 272116 | B | >5% | * |
| Sweden | 94283 | 1661484 | 5.67 | 17.62 | 583.0 | 164256 | B | >5% | -3.0 |
| Switzerland | 54384 | 1391498 | 3.90 | 25.58 | 239.0 | 160484 | B | 1-4.99% | -30.0 |
| Taiwan | 515 | 93914 | 0.54 | 182.35 | 0.3 | 3941 | A | <1.0% | -16.5 |
| Tunisia | 19721 | 238671 | 8.26 | 12.10 | 23.0 | 20142 | C | >5% | * |
| Turkey | 321512 | 10608664 | 3.03 | 32.99 | 98.0 | 125447 | C | 1-4.99% | -10.0 |
| U. Kingdom | 467146 | 25048460 | 1.86 | 53.62 | 622.0 | 368487 | B | 1-4.99% | -14.5 |
| Uruguay | 2026 | 238916 | 0.84 | 117.92 | 14.0 | 68717 | A | <1.0% | -0.6 |
| USA | 7507524 | 108562372 | 6.91 | 14.46 | 642.0 | 327494 | B | >5% | * |
| Vietnam | 1096 | 1009145 | 0.10 | 920.75 | 0.3 | 10344 | A | <1.0% | -16.3 |

**a**: cumulative data; **b**: Test positivity (%); **c**: Tests/case ratio; **d**: number of COVID-19 related deaths or tests performed per million inhabitants; **e**: country classification according to patterns exhibited by tests/case ratio and deaths/million inhabitants; **f**: country classification according to test positivity percentage; **g**: variation in the Gross Domestic Product of the second quarter of 2020.
